# Mindstep Mood and Cause Examination (MMCE): The Preferred Tool for Remote Digital Depression Screening

**DOI:** 10.1101/2024.08.07.24311602

**Authors:** M. Mahmud, N. Kuleindiren, S. Suddell, R. Rifkin-Zybutz, P. Sharma, T. Osunronbi, O. Pounds, H. Selim, A. Patchava, A. Lin, A. Alim-Marvasti

## Abstract

**Background:** Digital health technologies are increasingly being used to monitor, assess, and treat depressive symptoms in the community. However, many such technologies rely on screening tools which were originally designed for use in primary care clinics, such as the Patient Health Questionnaire (PHQ-9). These scales are symptom-focused and do not capture the wider experiences of the patient. We developed a new screen for assessing depressive symptoms in a digital setting. Named the Mindstep Mood and Cause Examination (MMCE), it was designed to replicate the predictive capabilities of the PHQ-9, while improving user experience and capturing broader determinants of mental health.

**Method:** This was a cross-sectional study, conducted fully remotely on Prolific. Participants (*n*=367) completed both the PHQ-9 and the MMCE, in a randomised order. Responses on the MMCE were examined for a range of psychometric properties, including: internal consistency, item selectivity, and convergence with PHQ-9 scores. User experience was assessed with a theory-led acceptability scale and compared across both mental health measures. Thematic analysis was used to analyse participants’ free text responses, describing their experience of completing the scales.

**Results:** The MMCE displayed good internal consistency and strong convergence with the PHQ-9 (*r* = 0.70), accounting for 49% of the variance in PHQ-9 scores. The MMCE also demonstrated robust predictive capability for the PHQ-9 using a moderate depression symptom cut-off of 10, with an Area Under Curve (AUC) of 0.84. In direct comparisons between the scales, 259 of 367 users (70.1%) preferred the MMCE and the MMCE outperformed the PHQ-9 in 8 out of 12 user experience categories.

**Conclusions:** The MMCE has demonstrated validity in predicting PHQ-9 scores and offers an improved user experience, while additionally encouraging the user to examine the underlying causes of their depressive symptoms. However, additional research is necessary to evaluate the MMCE in terms of repeated assessments for effective depression monitoring.

---

Depression affects over 300 million people globally and is a leading cause of disability and mortality worldwide [1]. The condition accounts for 10–20% of primary care visits, making it the second most common chronic condition seen by primary care physicians [2]. However, only half of these cases are identified by physicians, leaving many undiagnosed and untreated [2]. Barriers to depression diagnosis include inadequate treatment referral resources, limited access to mental health services, and an unequal geographical distribution of specialists [3, 4]. Digital health technologies, such as smartphone applications, can help overcome these barriers by facilitating depression screening remotely and at scale [5]. Users have shown high interest and adherence to smartphone applications for monitoring nearly all major psychiatric illnesses, with rates surpassing those of similar Internet-based interventions [6].

Many digital health platforms have repurposed pre-existing screening tools, originally evaluated for use in primary care clinics, to evaluate depression [7–9]. Commonly considered the gold-standard, the Patient Health Questionnaire (PHQ-9) is a depression rating scale that assesses symptoms of depression over the preceding two weeks and is widely used for screening, diagnosing, and monitoring of depression both in primary care and psychiatric clinics [10]. While the PHQ-9 has shown high sensitivity and specificity in detecting depression, it has received criticism for failing to reflect symptoms and experiences that are considered meaningful by patients [11]. Patients report struggling to ‘fit’ their experience of depression to the restrictive response options, and consequently fear their experiences are being misrepresented to clinicians [12]. In particular, by solely focusing on symptoms, the PHQ-9 fails to capture the most frequently reported challenges that patients bring to talking therapy, including work-related and relational issues, and therefore cannot determine patient-centred treatment priorities [13]. Perhaps of greater concern, some patients experience the PHQ-9 as distressing, particularly in regards to reporting self-harm and suicidal ideation, and subsequently underscore or omit answers to certain items [14]. These issues have contributed to clinicians expressing uncertainty regarding the use of the PHQ-9 as a replacement for clinical judgement [12].

These criticisms are amplified when the PHQ-9 is administered digitally in a remote setting, where patients are unable to provide any additional context to their condition. Furthermore, patients are at risk of experiencing distress caused by the PHQ-9 without the support of a clinician. Therefore, there is a pressing need to develop a new digital screening tool for depression, that can examine a wide range of patient experiences in a manner that is user-friendly, does not contain distressing items, and allows the patient to feel that their lived experience is understood. This is crucial as previous research has highlighted enhanced recovery efforts when patients feel understood by their GP [15]. An ideal digital tool should also maintain patient engagement which can be sustained by validation of patient experience and evidence shows that patient engagement decreases when such initial experiences do not meet these needs [16]. Using such a tool would allow digital health platforms to develop patient-centred treatment plans and support users in adhering to them.

Mindstep is a smartphone application designed to collect medical information and deliver personalised programmes focused on brain and mental health [17, 18]. A highly popular pathway within the app focuses on the monitoring of mild to moderate depressive symptoms that are endorsed by users. Signposting occurs for those who present with current risk to self, since this falls outside the scope of care offered. A tailored care plan can be undertaken over the course of several weeks, which has been shown as accessible for various ages and cognitive abilities [17, 18]. In addition to assessing symptom burden, it is crucial to understand the users’ wider experience of their depression to tailor this care plan appropriately. With this goal in mind, we have developed a novel depression screening tool, consisting of a set of ‘check all that apply’ questions, that examines not just symptom burden but also possible underlying drivers of mild to moderate depression, including questions about self and the world, values, money, relationships, and past trauma [19–23]. We called this screen the Mindstep Mood and Cause Examination (MMCE) (**Table 1**).

**Table 1.** Mindstep Mood and Cause Examination (MMCE).

| **Items and Responses** |
| --- |
| **Let's start with how you feel on the inside. Tap all points that feel true.**<br>• I often struggle to feel pleasure or joy<br>• I am often bothered by how low I feel<br>• I often lack interest in anything<br>• I often find I don't like myself<br>• None apply to me |
| **Moving onto the theme of 'work' - tap all that may apply to you.**<br>• I often worry about money<br>• My work situation is unstable<br>• I control my own time at work<br>• I find little joy in my work<br>• None apply to me |
| **In a similar vein, let's think about your values. Select all that apply.**<br>• I struggle to know what I want from life<br>• I pursue things that don't make me happy<br>• I've lost touch with what truly fulfils me<br>• My decisions don't reflect my inner values<br>• None apply to me |
| **Now onto people. As before, tap any of the following that you feel applies.**<br>• I often feel lonely<br>• I don't feel a sense of belonging<br>• I find people struggle to understand me<br>• Socialising is more stressful than fun<br>• None apply to me |
| **Ok, now let's reflect on your past. Please tap any that you feel apply.**<br>• I feel I have unresolved childhood trauma<br>• I often worry about things I have done<br>• I avoid discussing my childhood<br>• I worry a lot about repeating mistakes<br>• None apply to me |
| **Let's now think about the present. Select all that apply.**<br>• I feel undervalued by those around me<br>• My opinions/feelings are often dismissed<br>• I often feel inferior to others<br>• Others have it much better than I do<br>• None apply to me |
| **And lastly, let's focus on the future. Select all that apply.**<br>• I often feel hopeless about the future<br>• I often feel anxious about the future<br>• The uncertainty of the world worries me<br>• I feel the world is becoming more bleak<br>• None apply to me |

The items within the MMCE were developed following a literature review of common risk factors for depression. Such factors are multifactorial and vary across diverse populations and individual experiences, including work-related stress, financial hardships, loneliness, low self-esteem, adverse childhood experiences, and pessimistic future expectations [20–25]. Additionally, living in accordance with one’s values can significantly alleviate depressive symptoms [19]. When developing the MMCE, we designed questions and responses that capture these key drivers of depression, whilst also considering our data on the causes of low mood which has been collected from the users of Mindstep. We developed the questionnaire within a theoretical framework that is consistent with current best practice guidance to offer utility in using the questionnaire in conjunction with targeted first-line psychological interventions [26].

This study aimed to assess the validity, reliability, and acceptability of the novel MMCE scale for digital depression screening. In particular, we examine the psychometric properties of the MMCE in reference to the PHQ-9, to determine whether it can be used to predict established cut-offs of depressive symptoms. We then compare participants’ experiences of completing both scales to identify which scale is preferred by patients based on several usability features, including ease of use, language and tone, and relevance to their mental health experiences. Additionally, we collected qualitative feedback on both questionnaires.

## Method

### Study Design and Participants

Ethical approval for studies involving Mindstep data was originally obtained from the NHS Health Research Authority (West Midlands – Solihull Research Ethics Committee; REC reference – 21/WM/0202) on 16/09/21. This data was collected as post-market research for the purpose of product refinement. Informed consent was obtained from participants prior to their involvement in this study.

This study had a cross-sectional design with convenience sampling using Prolific, an online crowdsourcing platform, to recruit anonymous study participants, aged 18 to 80 years, who had a prior history of either anxiety or depression [27, 28]. Data collection occurred over three days, from 7 to 9 February 2024. Participants were administered both the MMCE and the PHQ-9 digitally and without supervision. Each participant accessed the questionnaire anonymously through a secure online platform, ensuring privacy and standardisation in the administration process. The participants were instructed to complete both questionnaires in a single session, allowing for a direct comparison of their responses between the two questionnaires. The order of the questionnaires was randomised across participants.

The PHQ-9 is a nine-item questionnaire where users indicate their endorsement of each item on a 0-3 Likert scale ranging from ‘not at all’ to ‘nearly every day’ [10]. The MMCE, by contrast, is a scale assessing seven domains of functioning (*symptoms, relationships, values, work, past, present, future*) in which users are asked to endorse any number of 4 statements within each domain (**Table 1**). A point is given for each answer selected, with increasing points assumed to indicate greater depression severity.

The entire survey consisted of socio-demographic questions (gender, ethnicity, age, employment status, educational history, and mental health history), the PHQ-9 and MMCE scales, and quantitative feedback on the acceptability of the PHQ-9 and MMCE scales. The participants’ data was anonymous, and the participants were compensated financially through their Prolific ID which cannot be linked to any personal information.

### Statistical Analysis

Our power calculation, performed using G*Power (version 3.1), recommended a minimum sample size of 84 based on the following parameters: two-tailed correlation bivariate normal model, effect size = 0.30, alpha level = 0.05, and power = 0.80.

All statistical analyses were performed in Python (v3.9.16), namely using the modules SciPy (1.7.3), statsmodels (v0.13.5), and scikit-learn (v1.5.1). Where applicable, Q-Q plots were used to examine the normality of data. Descriptive statistics were presented as mean and standard deviation for continuous variables and absolute count and percentages for categorical variables. In the interpretation of the linear regression models, p-values less than a Bonferroni-corrected significance level of 0.05/16=.003 were considered statistically significant.

The following psychometric properties were used to evaluate the MMCE questionnaire:

#### Floor and ceiling effects

We calculated the percentage of patients with the highest and lowest scores for the global MMCE scale and each MMCE subscale. Floor and ceiling effects were considered present if >15% of patients achieved the lowest (0) or highest score (4) within a subscale, or the lowest (0) or highest score (28) within the global scale, respectively [29].

#### Internal Consistency and Item Selectivity

We determined Cronbach’s alpha (α) for the global MMCE scale and each subscale. Based on Cronbach’s alpha, internal consistency was classified as either: excellent (α > 0.9), good (α = 0.8–0.9), acceptable (α = 0.7–0.8), questionable (α = 0.6–0.7), poor (α = 0.5–0.6), or unacceptable (α < 0.5) [30].

To determine item selectivity, we correlated each item with the global MMCE score, minus the value of the respective item. We also examined the correlation of the global score with each subscale score. As MMCE scores and items are not normally distributed, associations between global and subscale scores were analysed using the Spearman rank correlation coefficient, while associations between global MMCE scores (continuous) and individual items (binary) were analysed with point biserial correlations. The strength of the correlation was interpreted as strong (> 0.7), moderate (0.4 to 0.7) and weak (< 0.4) [31].

#### Convergent Validity

Convergent validity of the global MMCE and PHQ-9 scores was analysed using the Spearman rank correlation coefficient. The strength of the correlation was interpreted as strong (r > 0.7), moderate (0.4 to 0.7) and weak (r < 0.4) [31].

#### Concurrent Validity

We further examined the validity of the MMCE using a criterion-related approach to determine how well it can predict commonly used thresholds on the PHQ-9. Specifically, we evaluated the MMCE’s ability to classify individuals who do or do not meet the traditional PHQ-9 threshold of 10 (indicating moderate depressive symptoms). The area under the ROC curve (AUC) was used as an indicator of the measure’s classification accuracy. The AUC values were interpreted as: excellent (AUC > 0.9), good (AUC = 0.8–0.9), acceptable (AUC = 0.7–0.8), poor (AUC = 0.6–0.7), and failed (AUC = 0.5–0.6) [32].

An optimal threshold value for the MMCE was determined using Youden’s J statistic [33]. Sensitivity, specificity, positive predictive value (PPV), and negative predictive value (NPV) were calculated using this threshold. Confidence intervals for these metrics were calculated using the bootstrap method with 1,000 samples.

As an exploratory analysis, we then conducted a series of supervised machine learning models to optimise our prediction of depression status from global MMCE scores. The models implemented included logistic regression, support vector classifier, random forest, XGBoost, and k-nearest neighbours. For each model we performed 5-fold cross validation, and we report the mean performance metrics in the supplementary material.

#### Acceptability

Acceptability of the MMCE and PHQ-9 questionnaires were assessed using a modified version of a theory-led acceptability scale with the addition of questions specific to questionnaire acceptability [34], resulting in a 12-item scale. Each item was rated between 1 (very unacceptable) and 5 (very acceptable) and combined to compute an overall acceptability score ranging from 12 to 60. We conducted a series of paired t-tests to compare the rating of the scales for each acceptability item.

To examine the acceptability of the scales across participant groups, we conducted sensitivity analyses using a) depression severity and b) sociodemographic (age, gender, education) variables. These variables were entered in logistic and linear regression models, predicting questionnaire preference (MMCE or PHQ) and MMCE acceptability total scores, respectively.

A final, free-text item was included to allow participants to provide qualitative feedback regarding their experience of the two questionnaires. Participants were prompted to indicate which questionnaire they preferred and why. These free text responses were analysed using thematic analysis to identify common themes and insights.

#### Relationship with Sociodemographic Variables

Finally, as an exploratory analysis, we conducted two multiple linear regression models to investigate the association between the participants’ sociodemographic (age, gender, highest level of education, employment status, and ethnicity, and current depression diagnosis) and the PHQ-9 and MMCE global scores.

## Results

### Participant Characteristics

We recruited 396 participants in total, 367 of whom supplied data for both the MMCE and PHQ-9. Of this analytical sample, 68.9% identified as women and 89% of white ethnicity. Most participants were younger than 45 years (66.8%). The percentage of participants with an active diagnosis of depression or anxiety were 35% and 37% respectively. The mean PHQ-9 and MMCE scores were 11.1 ± 6.2 and 14.0 ± 6.3, respectively (**Table 2**).

**Table 2.** Participant demographics of the analytical sample (n = 367).

| **Variable** | **Response** | ***n*** | **%** |
| --- | --- | --- | --- |
| **Age** | Under 25 | 41 | 11.2 |
|  | 25-34 | 119 | 32.4 |
|  | 35-44 | 85 | 23.2 |
|  | 45-54 | 62 | 16.9 |
|  | 55-64 | 48 | 13.1 |
|  | 65-74 | 11 | 3.0 |
|  | 75+ | 1 | 0.3 |
| **Gender** | Woman | 253 | 68.9 |
|  | Man | 110 | 30.0 |
|  | Non-binary | 4 | 1.1 |
| **Employment** | Full-time employment | 184 | 50.1 |
|  | Full-time education | 15 | 4.1 |
|  | Education and employment | 11 | 3.0 |
|  | Part-time employment | 74 | 20.2 |
|  | Part-time education | 2 | 0.5 |
|  | Not in education, training, or employment | 60 | 16.3 |
|  | Stay at home parent | 21 | 5.7 |
| **Highest Education** | Postgraduate degree | 49 | 13.4 |
|  | Undergraduate degree | 143 | 39.0 |
|  | A-Levels | 101 | 27.5 |
|  | GCSEs | 67 | 18.3 |
|  | None | 7 | 1.9 |
| **Ethnicity** | White (includes British, Irish, Gypsy, Irish Traveller, Roma) | 328 | 89.37 |
|  | Mixed or Multiple ethnic groups | 16 | 4.36 |
|  | Asian or Asian British (Includes Indian, Pakistani, Bangladeshi, Chinese or any other Asian background) | 13 | 3.54 |
|  | Black, Black British, Caribbean or African (includes any other Black background) | 8 | 2.18 |
|  | Other ethnic group | 2 | 0.54 |
| **Mental Health Diagnosis** | Anxiety | 134 | 36.5 |
|  | Depression | 127 | 34.6 |
|  |  | *M* | *SD* |
| **Depression Scales** | PHQ-9 | 11.1 | 6.2 |
|  | MMCE | 14.0 | 6.3 |

*Note.* PHQ-9: Patient Health Questionnaire; MMCE: Mindstep Mood and Cause Examination.

### Psychometric properties of MMCE

#### Floor and Ceiling Effects

The global MMCE score showed no floor (0.5%) or ceiling (0.0%) effects. The ‘symptom’ (30.3%), ‘people’ (25.6%), ‘future’ (28.9%) and ‘present’ (15.3%) subscales showed ceiling effects. The ‘values’ (20.4%) and ‘present’ (18.3%) subscales showed floor effects (**Supplementary Table S1**).

#### Internal Consistency and Item Selectivity

The internal consistency of the MMCE global score was good (α = .87). However, the internal consistency of the ‘symptom’ (α = .68) and ‘people’ (α = .61) subscales were questionable, while those of the ‘values’ (α = .56), ‘future’ (α = .60) and ‘present’ (α = .60) subscales were poor. The internal consistency of the ‘work’ (α = .22) and ‘past’ (α = .49) subscales were unacceptable.

Item selectivity generally showed moderate effect sizes for both the MMCE global score and the individual subscales. The point biserial correlation between the items and the global MMCE score (excluding the respective item) ranged from *r*_pb_ *=.*02 to *r*_pb_ = .59, with 20 out of 28 items showing at least moderate (*r*_pb_ > .40) associations. All were statistically significant (*p <.*001) except for ‘I control my own time at work’ (*r*_pb_ *=.*02, *p* = .723). The three items with the highest correlation coefficients were: ‘I don’t feel a sense of belonging’ (*r*_pb_ *= .*59, *p < .*001), ‘I often feel hopeless about the future’ (*r*_pb_ *= .*58, p < .001), and ‘I often feel lonely’ (*r*_pb_ *= .*54, *p < .*001). The three items with the lowest correlation coefficients were all found in the ‘work’ subscale: ‘I control my own time at work’ (*r*_pb_ *= .*02, *p* = .723), ‘My work situation is unstable’ (*r*_pb_ *= .*23, *p < .*001), and ‘I find little joy in my work’ (*r*_pb_ *= .*23, *p < .*001).

There was strong correlation between the MMCE scale and the subscales, with the exception of ‘past’ and ‘work’ subscales which showed moderate correlation: ‘people’ (*r = .*81), ‘symptom’ (*r = .*78), ‘present’ (*r = .*78), ‘values’ (*r = .*75), ‘future’ (*r = .*74), ‘past’ (*r = .*67), and ‘work’ (*r = .*44) (**Figure 1**).

**Figure 1.**
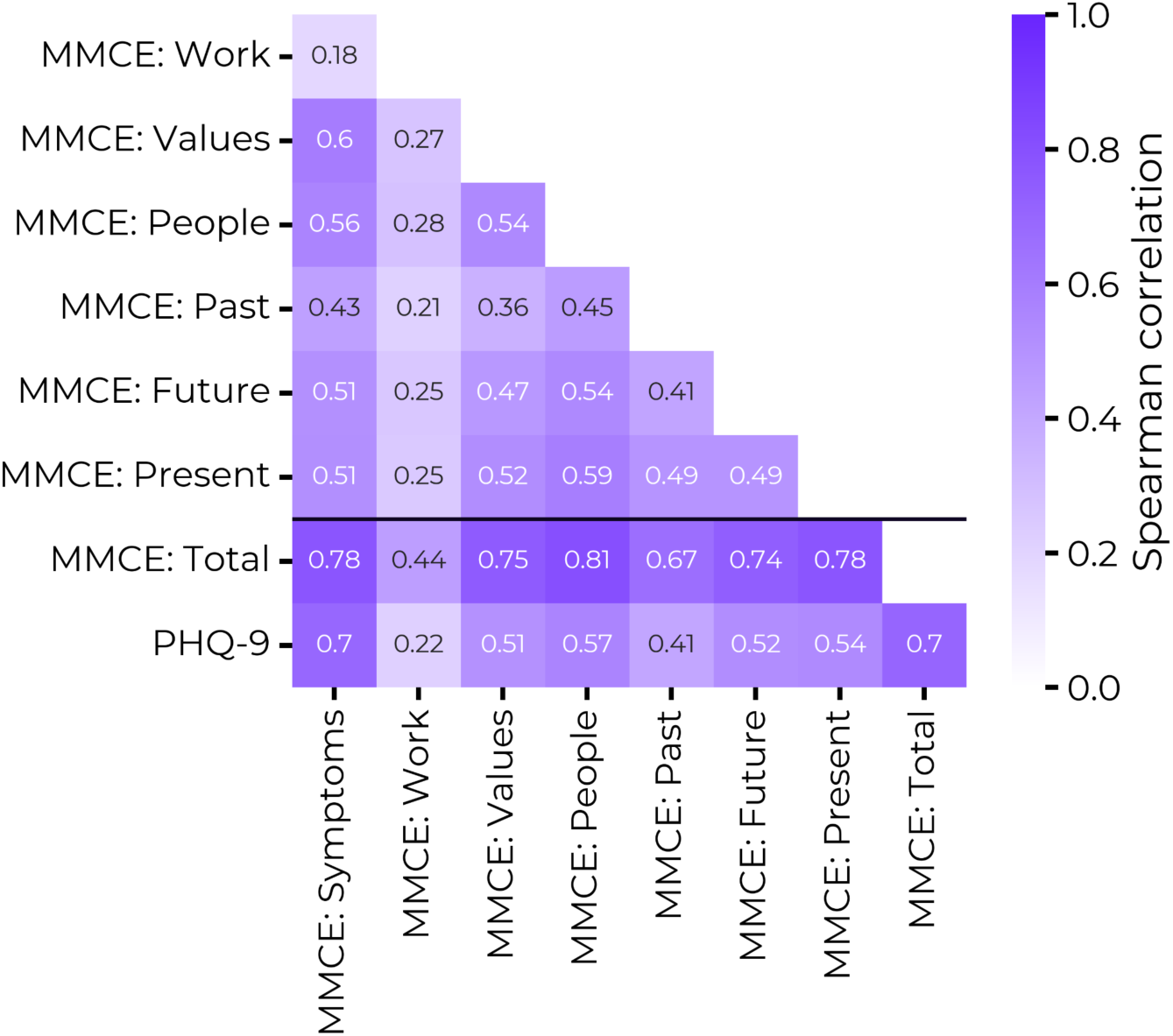
Pairwise correlations of MMCE subscales and PHQ-9 total scores.

#### Convergent Validity

There was a moderate positive correlation between the global MMCE and PHQ-9 scores, and this was statistically significant (*r* = .70, *p* < .001; **Figure 2**). Simple linear regression demonstrated that for a unit increase in the MMCE score, PHQ-9 score increases by 0.69, with MMCE score accounting for 49% variability in PHQ-9 score (*b* = 0.69, 95% CI = 0.61 to 0.76, *p* < .001, *R*² = .49).

**Figure 2.**
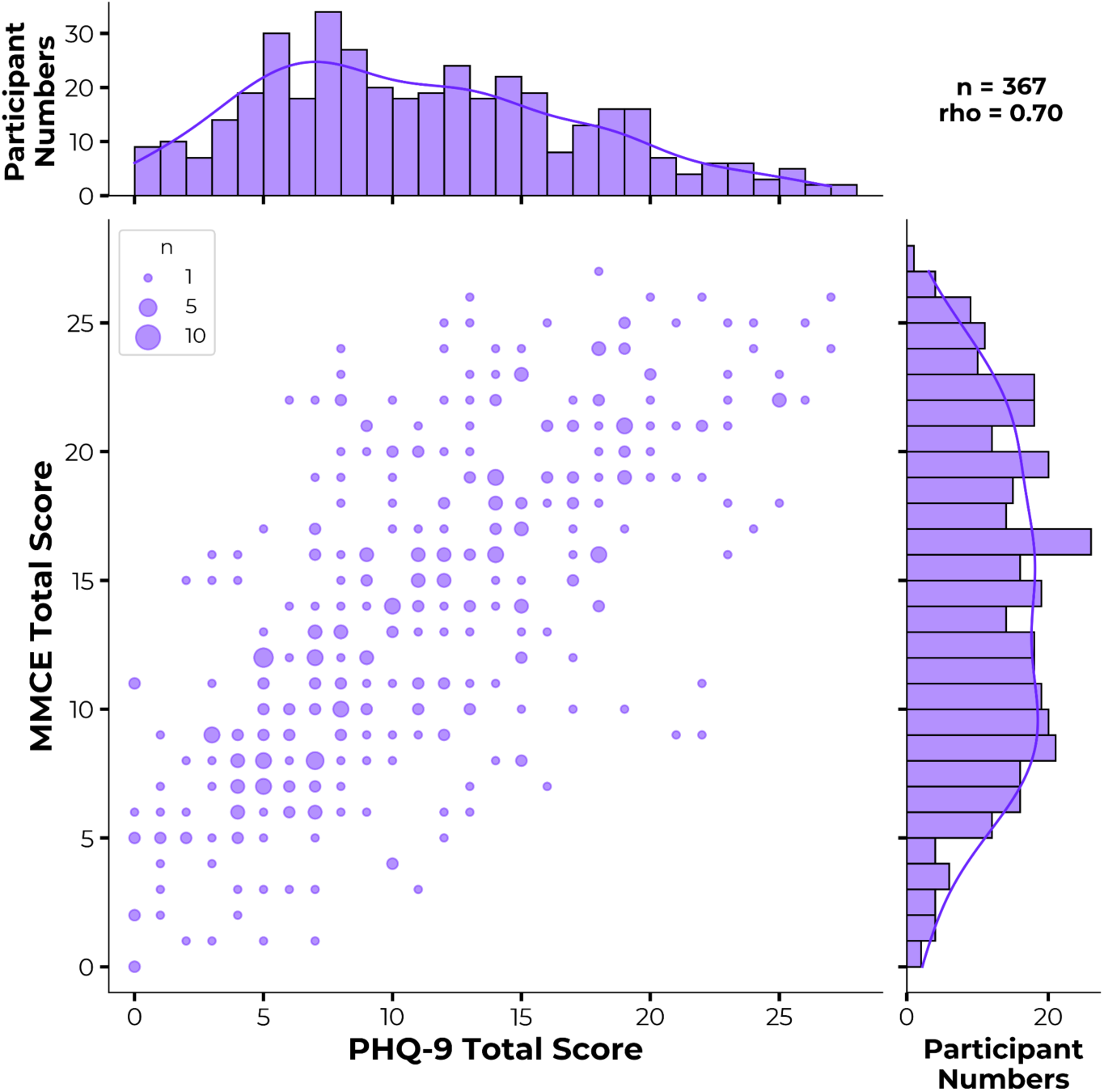
A bubble chart showing the correlation between users MMCE and PHQ-9 scores. The histograms above and to the right of the chart show the PHQ-9 and MMCE distributions respectively.

#### Concurrent Validity

Of the participants who had provided both an MMCE and PHQ-9 score, 202 participants (55%) had a PHQ-9 score ≥ 10, indicating moderate depressive symptoms, while 165 participants (45%) had a PHQ-9 score < 10, indicating no to mild depressive symptoms. Simple logistic regression analysis showed that a unit increase in the global MMCE score was associated with a 29% increase in the odds of a PHQ-9 score ≥10 (OR = 1.29, 95% CI = 1.23 to 1.36, *p* <.001).

The AUC of the MMCE global score in discriminating between these participants was .84 (95% CI = .80 to .88; **Figure 3**). The AUC value is in the ‘good’ range for discriminatory ability. At an optimal threshold value of MMCE >= 14, as calculated by Youden’s J statistic, the MMCE had a sensitivity of .79 (95% CI = .73 to .88) and a specificity of .79 (95% CI = .71 to .86). The positive predictive value was .82 (95% CI = .76 to .88) and the negative predictive value was .75 (95% CI = .69 to .83). A comparison of supervised classification models found comparable performance metrics across all models (**Supplementary Table S2**).

**Figure 3.**
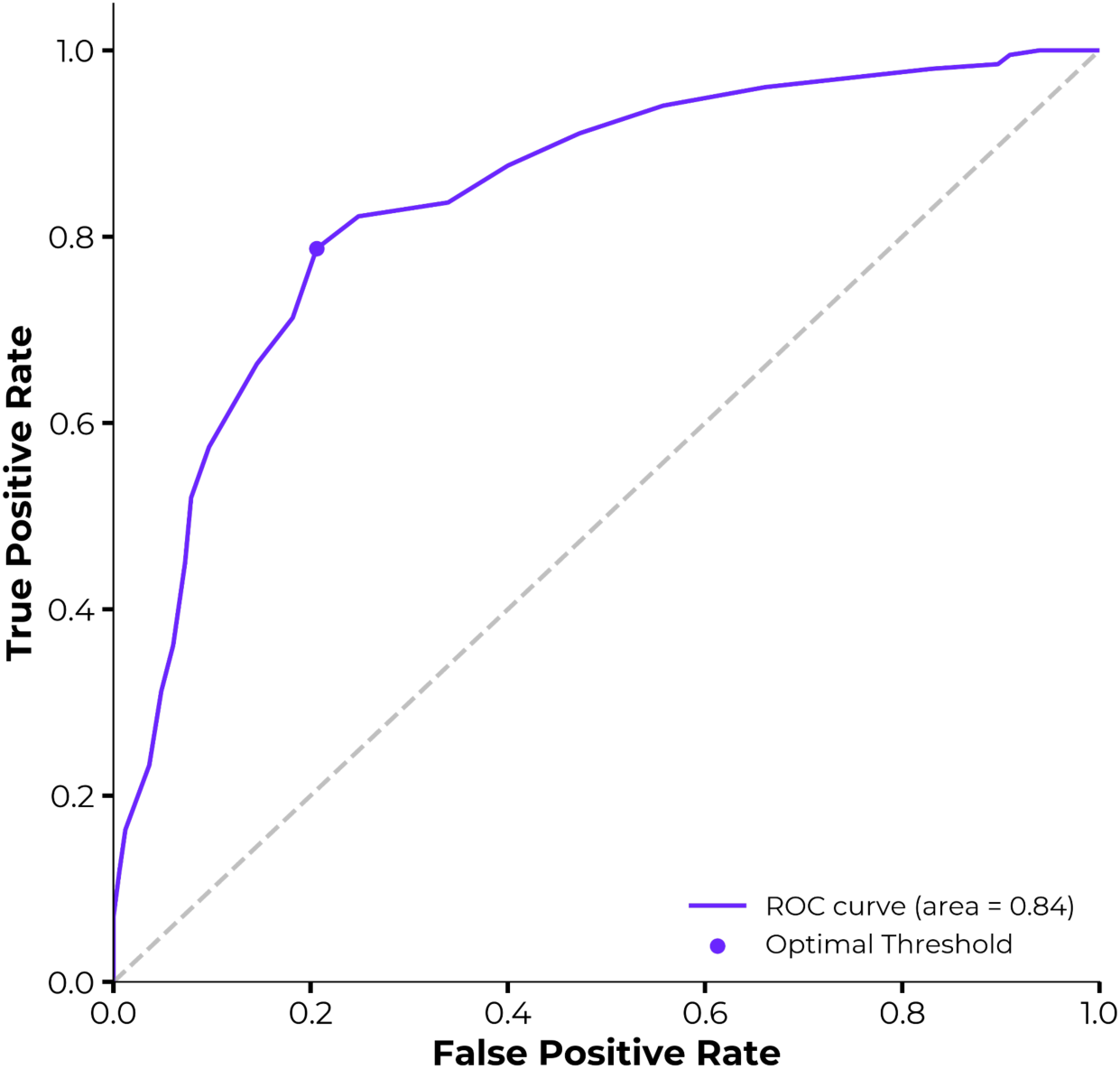
Receiving operating characteristic (ROC) curve for the prediction of PHQ-9 >= 10 using global MMCE score.

### Association with sociodemographic variables

Multiple linear regression models were built using all the sociodemographic variables (age, gender, highest level of education, employment status, and ethnicity) to predict PHQ-9 and MMCE scores. After Bonferroni correction (*p < .*003) in the PHQ-9 model, only ‘Not in education or employment’ (*b* = 4.49, 95% CI = 2.63 to 6.34, *p < .*001) was statistically significant — indicating that those with no occupation reported greater depressive symptoms (see **Supplementary Table S4**). A corresponding association was found for the MMCE global score (*b* = 3.08, 95% CI = 1.16 to 4.99, *p* = .002). In addition, age was negatively associated with MMCE global score (*b* = −1.03, 95% CI = −1.56 to −0.50, *p < .*001), indicating older participants scored lower.

### Acceptability of MMCE and PHQ-9

A paired t-test comparing overall scores on the acceptability scale showed that on average, the MMCE was rated 2.6 points higher than the PHQ-9 score (*t* = 5.87, *p* < .001). 259 of 367 users (70.1%) reported that they preferred the MMCE over PHQ-9 for digital screening of depression.

Examining individual items of the acceptability scale (**Figure 4**), both the PHQ-9 and MMCE questionnaires were generally regarded as relevant, positive, acceptable, requiring little effort, having a compassionate language and tone, and people generally liked them and would use them again. Both questionnaires received lower endorsement in terms of stimulating a deeper understanding of mental health, helping patients get to the core of their mental health, encouraging engagement with care, thoroughly evaluating mental health, and informing care decisions.

**Figure 4.**
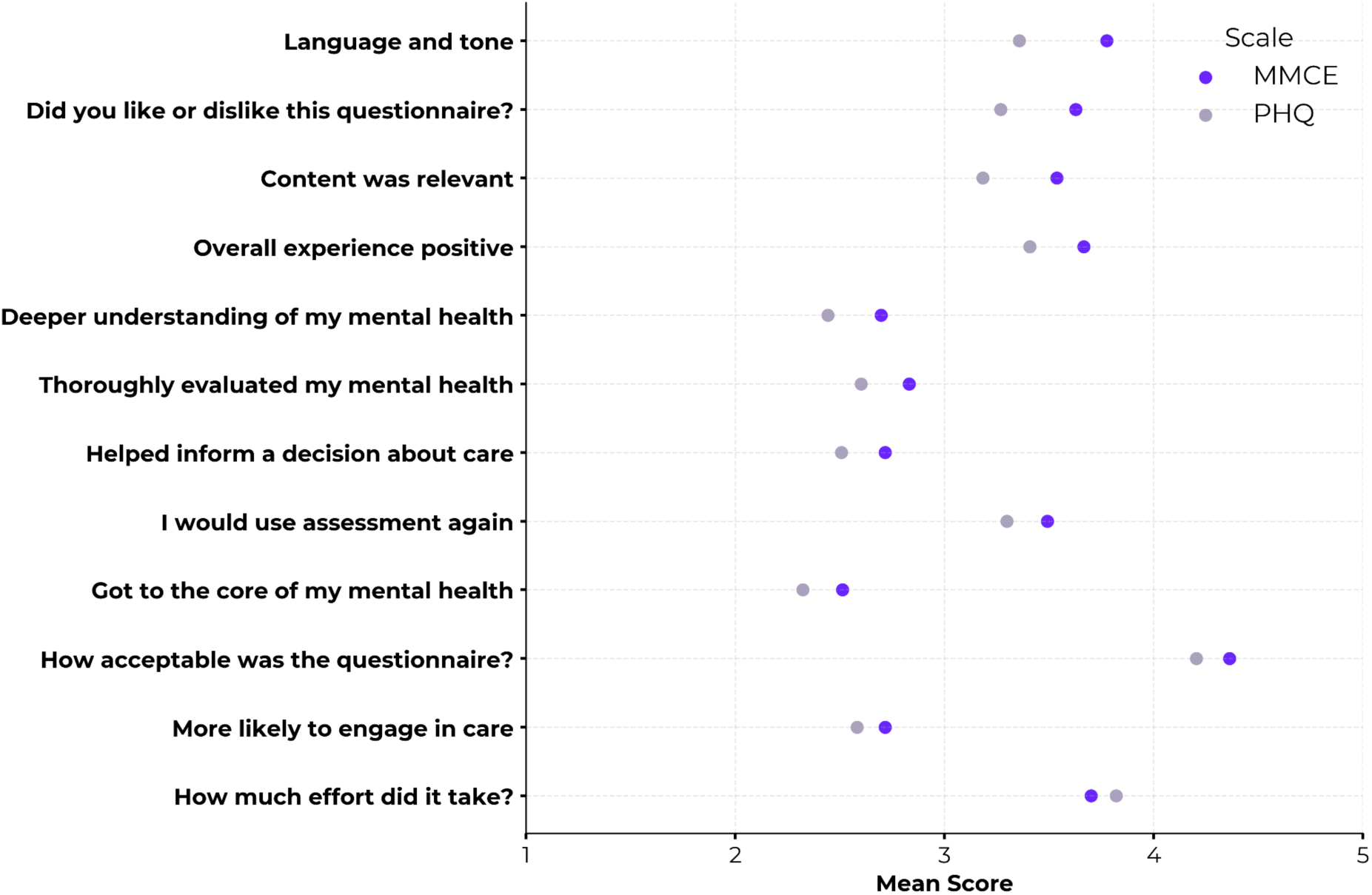
Mean Likert scores on each user acceptability, per mental health measure. All item responses ranged from 1 (*strongly disagree/high effort/completely unacceptable)* to 5 (*strongly agree/low effort/completely acceptable*).

The comparison of mean Likert scores for each item in the acceptability scale revealed that the MMCE significantly outperformed the PHQ-9 in 8 out of 12 items (Bonferonni adjusted for multiple comparisons). In particular, the MMCE was considered to have a far more compassionate language and tone, was significantly preferred over the PHQ-9, and was considered to include items of greater relevance to the patient’s experience of their depression than the PHQ-9. There were no statistically significant differences for the following 3 items: ‘*More likely to engage in care*’, ‘*Got to the core of my mental health*’, and ‘*I would use this assessment again*’ (**Figure 5, Supplementary Table S3**). On average the PHQ-9 was considered less effortful than the MMCE, however this was not significant at the Bonferroni-corrected threshold.

**Figure 5.**
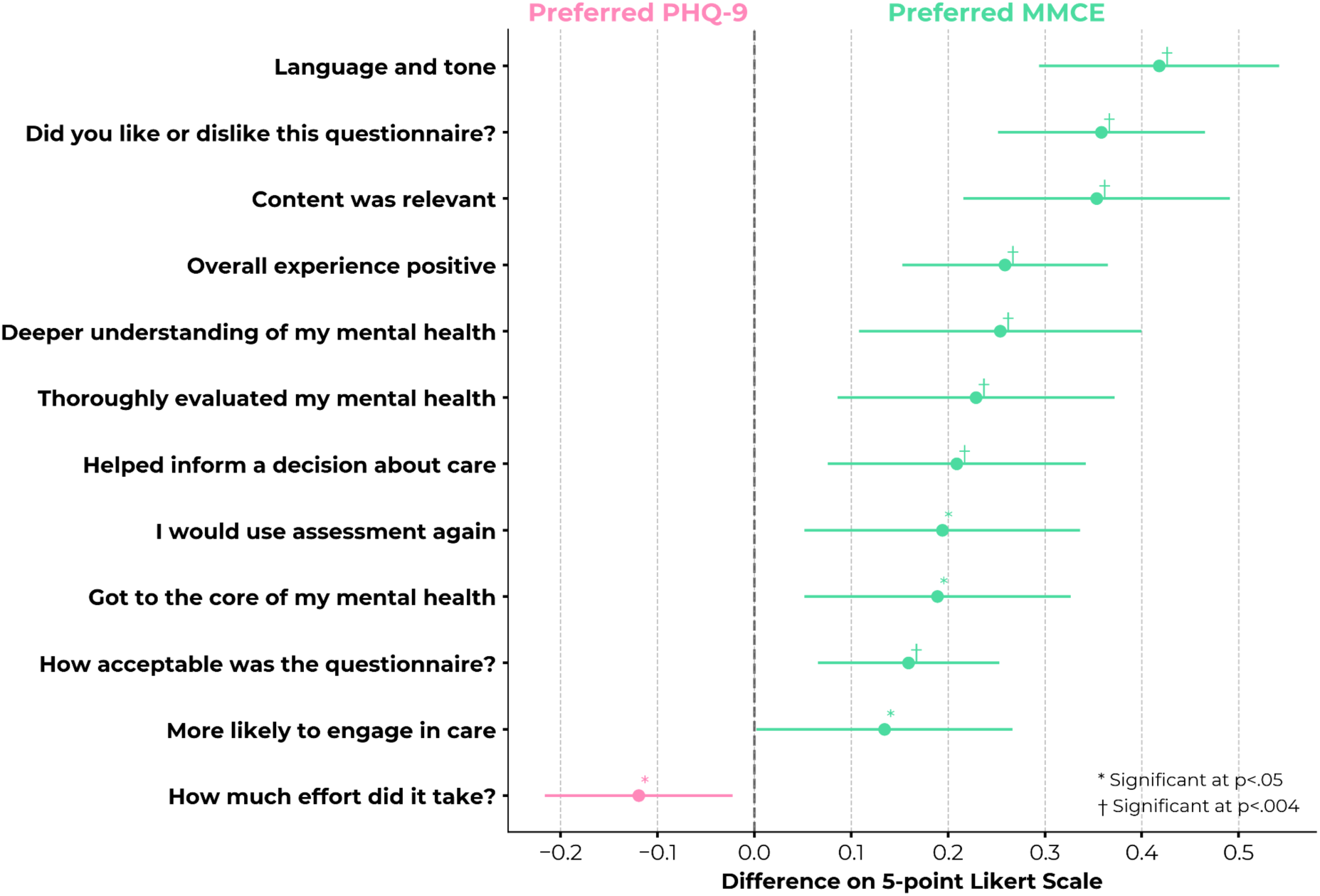
Forest plot showing the difference in mean Likert score between MMCE and PHQ-9 across 12 acceptability metrics. Symbols indicate significance at traditional *p<*.05 threshold (*) and *p<*.004 Bonferroni-corrected threshold (†).

To verify that the MMCE was acceptable to a wide range of user groups, we first examined user preferences across a range of depression severities, as determined by PHQ-9 score (**Figure 6**). All severity groups showed a preference for the MMCE over the PHQ-9, with this trend being strongest amongst users who reported no or mild depressive symptoms. A linear regression model, predicting global MMCE acceptability from PHQ-9 score, confirmed that there was no significant relationship between depression severity and MMCE acceptability score (*b =* 0.05, 95% CI = −0.11 to 0.21, *p* = .548). This indicates that the MMCE is deemed highly acceptable to participants across the full range of depressive symptom severity, and is particularly preferred by those with subclinical symptoms, who are likely to form a significant proportion of digital health users.

**Figure 6.**
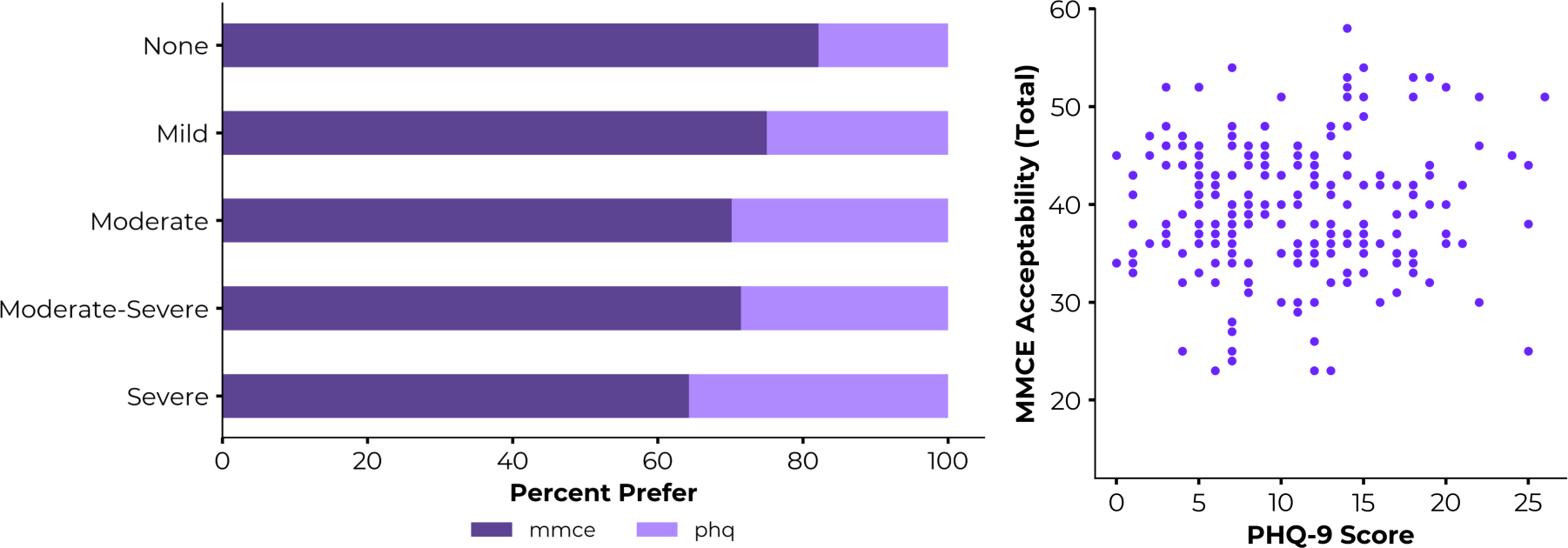
Examining the acceptability of the Mindstep Mood and Cause Examination (MMCE) across levels of depression severity, as measured by the PHQ-9. **Left:** The majority of users preferred the MMCE over the PHQ-9, irrespective of depression severity. **Right:** No relationship between PHQ-9 score and overall MMCE acceptability rating.

When examining user acceptability by sociodemographic factors, the MMCE was consistently rated as more acceptable on the 12-item scale, across all gender and age groups (**Figure 7**). In a logistic regression model predicting scale preference, we found no statistically significant effects of age (OR = 0.86, *p* = .097), gender (OR_male_ = 0.71, *p* = .171) nor highest level of education (compared to GCSE: OR_A-level_ = 1.18, OR_Undergrad_ = 1.15, OR_Postgrad_ = 0.90, all *p* > .600) in the likelihood of preferring the MMCE over PHQ-9. This suggests that the MMCE was consistently preferred over the PHQ-9 by participants from a broad range of demographic groups.

**Figure 7.**
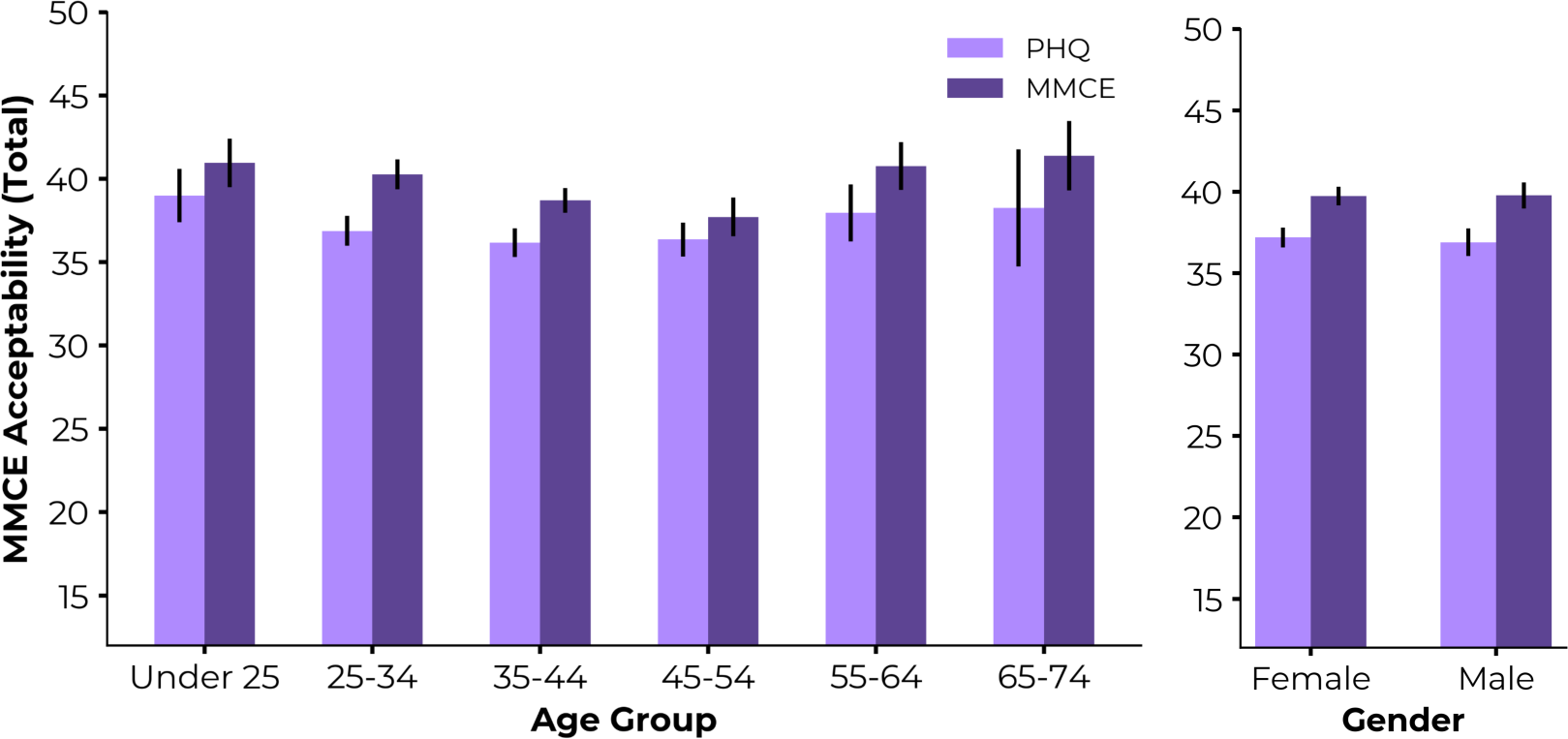
Examining the acceptability of the Mindstep Mood and Cause Examination (MMCE) across age and gender groups. The MMCE was consistently rated as more acceptable than the PHQ, irrespective of sociodemographic group.

Finally, to examine aspects of user acceptability that contributed to participants preferring the MMCE over the PHQ-9, we conducted a series of independent t-tests to compare the two preference groups (MMCE versus PHQ-9) on their responses to the 12 acceptability items. Significant differences between these two groups were identified for the items: *“thoroughly evaluated my mental health”* and *“deeper understanding of my mental health”.* Participants who preferred the MMCE tended to consider it as more thorough in evaluating their mental health (*t* = 3.18, *p* = .002) and felt the MMCE provided a deeper understanding of their mental health (*t* = 3.16, *p* = .002), than participants who preferred the PHQ-9.

### Thematic analysis

At the end of the survey, all participants were asked to reflect on the two questionnaires through free text responses. A total of 334 participants provided responses, which were used for thematic analysis. These responses were read by a member of the research team who identified initial codes, before grouping and refining these codes into broader themes. This process identified the themes of *emotional tone*, *relatability*, and *comprehensiveness*.

#### Emotional Tone

The MMCE was described as using gentler language and a more compassionate tone, making it feel more approachable and empathetic to participants. Participants appreciated the MMCE for its sensitivity:

> *“The [MMCE] used much nicer language which didn’t feel judgemental.”*

> (Female, 25-34)

> *“The [MMCE] felt closer to talking to a person.”*

> (Female, Under 25)

This contrasted with many participants’ description of the PHQ-9, which was described as having a more clinical and formal tone. One participant, a female aged 25-34, stated that *“the [PHQ-9] felt like a textbook and not compassionate”*, while another described completing the PHQ-9 as *“like answering to a robot”* (Female, Under 25). Others highlighted that some items of the PHQ-9 items were particularly challenging for them:

> *“[The question about] hurting myself made me feel uneasy.”*

> (Male, 45-54)

> *“[PHQ-9] uses terms which could be quite dark and upsetting.”*

> (Female, 55-64)

Only one participant described the tone of the MMCE negatively, making particular reference to the use of emojis in the presentation of the scale. They described this as “*almost patronising in a way*” (Female, Under 25). In contrast, one participant considered the MMCE as “*a more modern and personal approach*” (Female, 45-54).

#### Relatability

Participants often described the MMCE as more relatable, noting that the questions resonated with their personal experiences and made them reflect on their mental health. One participant, a male aged 25-34, expressed that “*the [MMCE] made me think a little more about my own mental health and being asked certain questions allowed me to learn more about myself*”. Another participant, a female under 25, found the MMCE to be “*more person-centred*”. Participants reported that the MMCE was more relevant to their lives, making it feel more tailored:

> *“I liked the [MMCE], I felt like I was able to identify more to it, particularly that it delved deeper”*

> (Male, 35-44)

> *“The [MMCE] felt easier to explain what was going through my head in the last couple weeks”*

> (Female, 25-34)

Conversely, the PHQ-9 was described as lacking detail and detached. One participant, male aged 25-34, stated that “*the [PHQ-9] was vague and didn’t help me assess my own mental health*”, while another participant mentioned how the PHQ-9 “*felt impersonal*” (Female, 25-34). Moreover, one participant, female aged 25-34, highlighted that the “*[PHQ-9] made it feel like if you weren’t suicidal then your problem isn’t that bad or as valid*”.

Notably, one participant reported that they preferred the MMCE, but emphasised the need for more options in certain scales to enhance inclusivity:

> *“I found the [MMCE] better but it didn’t have enough options for those in insecure situations due to their physical health. Didn’t have a self-employed option”*

> (Male, 25-34)

#### Comprehensiveness

MMCE was reported to be thorough, delving into greater depth regarding the participants’ mental health and focusing on underlying contributing factors:

> *“The [MMCE] actually hones in on what specifically makes you unhappy”*

> (Male, 25-34)

> *“The [MMCE] had a better understanding of daily moods and feelings”*

> (Male 35-44)

In contrast, the PHQ-9 was described as brief and being unable to provide additional insight to the participants. One participant, female aged 25-34, stated that “*the [PHQ-9] only explored certain aspects of mental health, e.g. going through the depression checklist but didn’t explore much more*” and another added that “*my main bug with it is that it focuses on frequency and ignores intensity of feelings*” (Female, 65-74). Further responses noted the brevity of the PHQ-9:

> *“The [PHQ-9] was too short, not detailed enough to assess mental health”*

> (Female, 35-44)

> *“The [PHQ-9] seemed too short”*

> (Female, 35-44)

## Discussion

In this report we examined the reliability, validity, and acceptability of a novel scale, the Mindstep Mood and Cause Examination (MMCE), designed specifically for the purpose of conducting depression screening in a digital setting. Our examination of the psychometric properties suggests that the MMCE is a valid scale with good convergence with the PHQ-9, the current gold-standard for assessing depressive symptoms in a primary care setting [10]. Crucially, as the MMCE will be delivered remotely and without clinical supervision, it was necessary to build upon the PHQ-9 in several key ways. Firstly, the MMCE needed to be user friendly, with a compassionate language and tone, and without the inclusion of items pertaining to self-harm. Secondly, the MMCE needed to capture a wider range of patient experiences than the symptom-focussed PHQ-9, as patients are often unable to provide additional context in a digital setting. Finally, the scale should encourage patients to self-reflect on the potential causes of their low mood, in order to initiate their recovery. Results from our acceptability analyses suggest the MMCE has achieved this, with participants considering it as having a more compassionate tone, more relevant content, and inspiring a deeper understanding of their mental health than the PHQ-9. Users preferred using the MMCE to the PHQ-9, and, in qualitative responses, reported it was more relevant to their lived experience.

The MMCE demonstrated strong convergent validity with the PHQ-9, and robust predictive capability in identifying moderate depression cases (PHQ-9 score > 10). This indicates the MMCE is effective in measuring depression symptoms in a manner consistent with the established gold-standard. We also found comparable associations with sociodemographic variables across both scales, with previously-identified factors such as age and employment status being significantly associated with depression severity [35]. The lack of floor or ceiling effects in the MMCE global score demonstrates that the scale captures a range of severity levels. Taken together, these analyses suggest that the MMCE is a valid tool for measuring depressive symptoms at scale and in a remote community setting.

We observed good internal consistency within the MMCE and, overall, results from the item selectivity analysis suggested moderate correlations between each individual item and the global score. This indicates that most items within the MMCE effectively contribute to a coherent measurement of the experience of depression. A notable exception was for items concerning work-related experiences, with both the individual items and the work-related subscore showing poor consistency with the overall score. This theme primarily includes items pertaining to circumstantial factors (such as being able to control one’s time at work) rather than the internal mood states captured within the remaining themes. This low consistency could in-part be driven by the fact that one of the items was positively phrased, in contrast to all of the others. Given this divergence, this theme requires further refinement. One approach could be to eliminate the work-related theme entirely to maintain the questionnaire’s focus on internal psychological states. However, it is important to note that various working conditions have been shown to put individuals at a greater risk of developing depression, and therefore it may be beneficial to instead focus on specific work-related factors such as working greater than 52 hours a week or shift work in the MMCE to improve the relevancy of the ‘work’ subscale [36, 37]. Furthermore, Mindstep is designed to produce targeted care plans for the user, and the domain of work represents a considerable area for intervention in which large mental health gains could be made.

In terms of user acceptability, the MMCE was consistently rated as a more positive user experience than the PHQ-9, with 7 out of 10 people preferring it to the PHQ-9. This pattern remained consistent across a range of sociodemographic groups and levels of depression severity, indicating that the MMCE has wide acceptability. While participants regarded the experience of completing both questionnaires as positive, they endorsed preferring the MMCE in 8 out of 12 acceptability domains. The PHQ-9 outperformed the MMCE in only one domain– perceived effort–as participants regarded the MMCE as more effortful. This is likely due to the extra consideration needed from the user when reflecting on which experiences relate to them – which, arguably, is one of the intentions behind this design. Crucially, participants reported that the MMCE encouraged a deeper understanding and provided a more thorough evaluation of their mental health than the PHQ-9. These aspects of user acceptability are particularly important in a digital setting, where patients are often unable to interact with a clinician. In the context of traditional GP consultations, patients often report the very act of being understood and listened to helps make sense of their problems, defines relevant outcomes, and supports their efforts to recover [15, 38]. A tool such as the MMCE, that allows patients to feel their experiences are captured even in a remote setting, could be a catalyst to aid with goal-setting and self-efficacy that will help promote recovery and potentially increase engagement with care.

Thematic analysis revealed several key insights regarding the strengths and limitations of the MMCE and the PHQ-9 when administered in a digital setting. Firstly, the MMCE’s friendly and compassionate language may be able to encourage patients to engage more readily with the tool, promoting regular use, and fostering a more supportive screening environment. However, this must be balanced with maintaining a professional tone to avoid making patients feel patronised. In contrast, the PHQ-9’s more clinical and dark tone was reported to be distressing by several patients, which is a finding supported by previous research [14]. This could deter patients from fully completing the PHQ-9 or lead them to omit answers to certain questions, thereby complicating the diagnostic process. In terms of relatability, participants expressed that the MMCE prompted deeper reflection on their mental health, making the experience more personal and meaningful. This contrasts with the PHQ-9, which was perceived as impersonal and difficult to relate to unless participants were experiencing suicidal thoughts. Lastly, the comprehensiveness of the MMCE was noted as a significant strength, as it attempts to delve into potential causes of depression. Conversely, the PHQ-9 focuses solely on the frequency of certain symptoms which participants felt was insufficient for a holistic understanding of their mental state. Another study highlighted patient feedback from those who completed the PHQ-9 in a primary care setting, noting that the questionnaire’s focus on symptom frequency, rather than intensity, might not fully capture the risk associated with extreme states of distress [11].

### Limitations

Although the questionnaire attempts to contextualise and offer an understanding of user experiences of depression, above that of the PHQ, a limitation of the MMCE is that in trying to offer a questionnaire that is brief and focused on possible causes of depression, it is limited in capturing an exhaustive list of experiences and in offering wider sociocultural aspects of depression. In particular, it should be acknowledged that our sample is predominantly from a white ethnic background and therefore the MMCE may not be as applicable to individuals who are from non-Western and/or non-white backgrounds. Expanding the questionnaire to include questions about situational factors, such as grief and loss, marital issues and physical health comorbidities, would enhance its relevance and applicability for broader audiences. Such additions might also facilitate targeted interventions, like those addressing grief, by providing more nuanced insights into the contexts affecting individuals’ mental health. It is important to note that the data collected here is cross-sectional, and so it remains to be seen how the MMCE compares to the PHQ-9 in terms of longitudinal assessment of depression trajectories. Finally, while we excluded items pertaining to self-harm and suicide, given previous reports that these can be distressing, Mindstep has appropriate safeguarding and signposting in place for patients to ensure that their more critical needs are cared for in a timely manner.

### Future Directions

Next steps should consider how to refine the existing set of items to increase assessment depth and applicability to wider populations, whilst still considering ease of use. This may include redeveloping the items pertaining to work, while also adding domains relevant to a wider set of users. Furthermore, the MMCE should be evaluated as a tool to measure depressive symptom change overtime.

Wider future directions include developing and testing targeted interventions based on individual responses, following the completion of the questionnaire. Digital screening tools are highly desired to relieve strain on public healthcare services [39]. The MMCE delivered via the app could be applicable in various clinical settings, offering screening and early tailored support for those attending primary care settings or for those on waiting lists for clinician-delivered psychological intervention. Using the results of the MMCE to produce targeted care plans would maximise the clinical utility of the MMCE. For instance, NICE guidance recommends befriending and rehabilitation as possible interventions in depression, which may be useful for individuals selecting unemployment or loneliness as contributing factors to mental health difficulties [26]. Use of the MMCE, in comparison to PHQ-9, would enable us to direct users to more relevant resources, and the inclusion of bespoke responses may increase adherence to care plans.

### Conclusion

The MMCE has demonstrated strong convergent validity with the PHQ-9 and robust predictive capabilities for assessing depression severity. Moreover, the MMCE has been shown to be the patient preferred tool for remote digital depression screening due to its compassionate, comprehensive, and personal approach. Future challenges for the MMCE include offering comprehensive assessment of possible underlying factors in depression across a non-specific and diverse population, whilst maintaining ease and user experience. There are also challenges with digitally administered questionnaires in offering tailored feedback to support future directions for users. Overall, the study supports the MMCE’s utility as a digital depression screening tool, aimed at both symptom evaluation for diagnostic purposes and improving user experience through understanding of underlying causes.

## Data Availability

All data produced in the present study are available upon reasonable request to the authors.

