## Supplementary material for "Mindstep Mood and Cause Examination (MMCE): The Preferred Tool for Remote Digital Depression Screening": Table S1

**Table S1.** Floor and ceiling effects in the Mindstep Mood and Cause Examination (MMCE), per subscale.

|  | **Proportion with lowest score (%)** | **Proportion with highest score (%)** |
| --- | --- | --- |
| **Global** | 0.5 | 0.0 |
| **Symptom** | 11.7 | 30.3 |
| **Work** | 10.9 | 3.5 |
| **Values** | 20.4 | 7.4 |
| **People** | 10.6 | 25.6 |
| **Past** | 13.9 | 12.0 |
| **Future** | 6.5 | 28.9 |
| **Present** | 18.3 | 15.3 |
