## Supplementary material for "Mindstep Mood and Cause Examination (MMCE): The Preferred Tool for Remote Digital Depression Screening": Table S2

**Table S2.** Comparison of supervised classification models for predicting moderate depression status with global MMCE score.

|  | **AUC** | | **Accuracy** | | **Sensitivity** | | **Specificity** | |
| --- | --- | --- | --- | --- | --- | --- | --- | --- |
| **Model** | ***M*** | ***SD*** | ***M*** | ***SD*** | ***M*** | ***SD*** | ***M*** | ***SD*** |
| **Logistic Regression** | 0.85 | 0.04 | 0.79 | 0.04 | 0.82 | 0.05 | 0.75 | 0.10 |
| **Support Vector Classifier** | 0.84 | 0.05 | 0.79 | 0.05 | 0.85 | 0.05 | 0.72 | 0.12 |
| **Random Forest** | 0.82 | 0.04 | 0.74 | 0.06 | 0.76 | 0.09 | 0.72 | 0.11 |
| **XGBoost** | 0.82 | 0.06 | 0.76 | 0.08 | 0.78 | 0.09 | 0.73 | 0.12 |
| **K-Nearest Neighbours** | 0.79 | 0.04 | 0.74 | 0.06 | 0.78 | 0.08 | 0.69 | 0.12 |

*Note.* MMCE: Mindstep Mood and Cause Examination.

Moderate depression status defined as PHQ-9 score>10.
