## Supplementary material for "Mindstep Mood and Cause Examination (MMCE): The Preferred Tool for Remote Digital Depression Screening": Table S3

**Table S3.** Comparison of mean Likert scores for each item in the acceptability scale for MMCE and PHQ-9.

| **Item** | **MMCE Mean** | **PHQ-9 Mean** | ***p*** |
| --- | --- | --- | --- |
| Helped make a more informed decision about care? | 2.72 | 2.51 | .002 |
| Got to the core of my mental health | 2.51 | 2.32 | .007 |
| I would use assessment again | 3.49 | 3.30 | .008 |
| Overall Experience positive | 3.67 | 3.41 | <.001 |
| Content was relevant | 3.54 | 3.18 | <.001 |
| I am more likely to engage in care after this assessment | 2.72 | 2.58 | .047 |
| I have a deeper understanding of my mental health after this assessment | 2.70 | 2.44 | <.001 |
| Language and tone of assessment compassionate | 3.78 | 3.36 | <.001 |
| Thoroughly evaluated my mental health | 2.83 | 2.60 | .002 |
| How acceptable was the questionnaire to you? | 4.36 | 4.20 | <.001 |
| How much effort did it take to complete this questionnaire? | 3.70 | 3.82 | .016 |
| Did you like or dislike this questionnaire? | 3.63 | 3.27 | <.001 |

*Note.* PHQ-9: Patient Health Questionnaire; MMCE: Mindstep Mood and Cause Examination.

All items have a possible range of 1 (*Strongly Disagree*) to 5 (*Strongly Agree*).
