## Supplementary material for "Mindstep Mood and Cause Examination (MMCE): The Preferred Tool for Remote Digital Depression Screening": Table S4

**Table S4.** Linear regression models predicting the PHQ-9 and MMCE with sociodemographic variables.

|  | **PHQ-9** | | |  | **MMCE** | | |
| --- | --- | --- | --- | --- | --- | --- | --- |
|  | ***b*** | **95% CI** | ***p*** |  | ***b*** | **95% CI** | ***p*** |
| **Age** | -0.62 | -1.13, -0.10 | .019 |  | -1.03 | -1.56, -0.50 | <.001 |
| **Employment Status (ref: full-time employment)** |  |  |  |  |  |  |  |
| Education & employment | 2.02 | -1.78, 5.81 | .297 |  | 1.40 | -2.53, 5.33 | .483 |
| Full-time education or training | -0.8 | -4.17, 2.58 | .643 |  | -3.03 | -6.52, 0.47 | .089 |
| Part-time education or training | 2.12 | -6.31, 0.53 | .621 |  | -0.04 | -8.75, 8.66 | .992 |
| Part-time employment | 0.15 | -1.55, 1.86 | .859 |  | -0.08 | -1.85, 1.68 | .927 |
| Stay at home parent | 0.78 | -2.04, 3.60 | .585 |  | -0.28 | -3.20, 2.64 | .849 |
| Not in education, employment or training | 4.49 | 2.63, 6.34 | <.001 |  | 3.08 | 1.16, 4.99 | .002 |
| **Ethnicity (ref: white)** |  |  |  |  |  |  |  |
| Black | 2.07 | -2.21, 6.35 | .342 |  | 0.83 | -3.60, 5.25 | .714 |
| Mixed | 2.44 | -0.61, 5.49 | .117 |  | 2.59 | -0.57, 5.74 | .108 |
| Other | -0.55 | -9.11, 8.00 | .899 |  | -3.55 | -12.39, 5.30 | .431 |
| **Gender (ref: female)** |  |  |  |  |  |  |  |
| Male | -1.01 | -2.44, 0.41 | .163 |  | -0.72 | -2.19, 0.75 | .338 |
| Non-binary | 2.91 | -3.14, 8.95 | .345 |  | 0.98 | -5.28, 7.23 | .759 |
| **Education (ref: postgraduate)** |  |  |  |  |  |  |  |
| None | 2.78 | -2.05, 7.60 | .258 |  | 1.88 | -3.11, 6.87 | .458 |
| GCSE | -1.51 | -3.84, 0.81 | .202 |  | -0.47 | -2.87, 1.94 | .703 |
| A-Level | 0.54 | -1.58, 2.66 | .615 |  | 0.21 | -1.98, 2.40 | .850 |
| Undergrad | -0.04 | -2.01, 1.93 | .969 |  | 0.34 | -1.70, 2.37 | .743 |

*Note.* PHQ-9: Patient Health Questionnaire; MMCE: Mindstep Mood and Cause Examination.
